# Strongly Divergent Impact of Adherence Patterns on Efficacy of Colorectal Cancer Screening: The Need to Refine Adherence Statistics

**DOI:** 10.1101/2020.10.05.20206854

**Authors:** Thomas Heisser, Rafael Cardoso, Feng Guo, Tobias Moellers, Michael Hoffmeister, Hermann Brenner

## Abstract

**Objective:** The performance of colorectal cancer (CRC) screening programs depends on the adherence to screening offers. However, identical adherence levels may result from varying patterns of the population’s screening behavior. We quantified the effects of different adherence patterns on the long-term performance of CRC screening for annual fecal immunochemical testing (FIT) and screening colonoscopy at ten-year intervals.

**Design:** Using a multistate Markov model, we simulated scenarios where, while at the same overall adherence level, a certain proportion of the population adheres to all screening offers (‘selective’ adherence) or the entire population uses the screening offers at some point(s) of time, albeit not in the recommended frequency (‘sporadic’ adherence). Key outcomes for comparison were the numbers of prevented CRC deaths and prevented years of potential life lost (YPLL) after 50 simulated years.

**Results:** For screening with annual FIT at adherence levels of 10-50%, ratios of prevented CRC deaths (YPLL) resulting from a sporadic versus a selective pattern ranged from 1.9-5.0 (1.9-5.0) for men and from 1.8-4.1 (1.8-4.3) for women, i.e. up to 4-5 times more CRC deaths and YPLL were prevented when the population followed a sporadic instead of a selective adherence pattern. Comparisons of simulated scenarios for screening colonoscopy revealed similar patterns.

**Conclusion:** At the same overall level of adherence, ‘sporadic’ adherence patterns go along with much larger preventive effects than ‘selective’ adherence patterns. Screening programs should prioritize efforts to reach as many people as possible at least sporadically over efforts to maximize full adherence to repeat screening offers. Adherence statistics should be refined to better reflect ‘effective adherence’.

**What You Need to Know:** *BACKGROUND AND CONTEXT:* The evidence on long-term effects of different patterns of longitudinal adherence (e.g. consistent or sporadic uptake) to colorectal cancer screening offers is limited.

*NEW FINDINGS:* In a simulated hypothetical German population, at identical overall participation levels, large proportions of the population making sporadic use of screening offers were up to 4-5 times more beneficial to achieve sustained reductions of colorectal cancer mortality than small proportions of the population utilizing screening offers at the recommended frequency.

*LIMITATIONS:* This study is limited by model simplifying assumptions and uncertainties related to input parameters.

*IMPACT:* Efforts to increase screening uptake should be concentrated on groups of consistent non-responders, e.g. by low-threshold screening offerings, such as directly mailed stool tests. Adherence statistics should be refined to better reflect “effective adherence”.

*SHORT SUMMARY:* This simulation study demonstrates that commonly used adherence metrics for colorectal cancer screening do not sufficiently cover the effect of varying patterns of longitudinal adherence, which may considerably impact the long-term efficacy of screening programs.

## Introduction

It is widely accepted that screening for colorectal cancer (CRC) is a very effective and cost-effective approach to reduce CRC incidence and mortality.^1–4^ Screening exams most commonly recommended by expert panels and offered in screening programs around the world include fecal immunochemical tests (FITs) and screening colonoscopy. Recommended starting ages for screening in the general average risk population range between 45 and 60 years, with proposed screening intervals of either 1 or 2 years for FITs and 10 years for colonoscopy.^5,6^ While these screening strategies may be effective per se, the performance of a population-based screening program also depends, to a large extent, on the adherence to screening offers.

Adherence to screening on the population level is commonly quantified by the proportion of the population in the eligible age range that is up to date with screening, i.e., that has had a FIT within the past 1 or 2 years, or a colonoscopy within the past 10 years.^7–9^ These metrics, however, do not differentiate between situations in which a certain proportion of the population adheres to all screening offers and situations in which a large proportion of the population uses the screening offers at some point(s) of time, albeit not in the recommended frequency. For instance, an adherence pattern where half of the eligible population makes use of an annual FIT screening offer will yield the same overall uptake level as a pattern where the entire population makes use of the offer only every second year. Such hidden variations in utilization patterns could, however, result in considerable differences in the effectiveness of comparably designed screening programs, as the incremental benefit of screening is likely higher for first-time than for repeated uptake.^10^ Understanding long-term consequences of differential adherence patterns may therefore be highly relevant to health authorities seeking to design and optimize organized screening programs.

Although several studies have addressed coverage and participation of CRC screening offers, ^11–13^ the evidence on long-term effects of different adherence patterns is limited.^14^ We therefore performed a simulation study in order to quantify expected effects of FIT- and colonoscopy-based screening offers in terms of prevented CRC deaths and years of life lost (YPLL) due to CRC for two distinct patterns of adherence that would go along with seemingly equivalent adherence metrics.

## Methods

### Multistate Markov Model

For this analysis, we used a previously developed and validated multistate Markov model simulating effects of screening for CRC in a hypothetical German population. A documentation on the model’s structure and data sources used for its development is provided in **Supplementary Appendix 1a**, including overviews on all model parameters (**Appendix 1a, Tables 1-3**).

**Table 1.** Differences in Long-term Outcomes for Screening with Annual FIT from Ages 50-75 given Varying Patterns of Adherence Yielding Identical Adherence Levels

**Men**
| Selective adherence |  |  | Sporadic adherence |  |  | Ratio sporadic / selective |  |
| --- | --- | --- | --- | --- | --- | --- | --- |
| Scheme | Prevented CRC deaths | Prevented YPLL | Scheme | Prevented CRC deaths | Prevented YPLL | Prevented CRC deaths | Prevented YPLL |
| 100% annually | 5,531 (90%) | 62,043 (86%) | 100% annually | 5,531 (90%) | 62,043 (86%) | 1.0 | 1.0 |
| 50% annually | 2,765 (45%) | 31,021 (43%) | 100% every 2 years | 5,178 (84%) | 57,473 (80%) | 1.9 | 1.9 |
| 33% annually | 1,844 (30%) | 20,681 (29%) | 100% every 3 years | 4,766 (77%) | 52,493 (73%) | 2.6 | 2.5 |
| 25% annually | 1,383 (22%) | 15,511 (22%) | 100% every 4 years | 4,376 (71%) | 47,948 (67%) | 3.2 | 3.1 |
| 20% annually | 1,106 (18%) | 12,409 (17%) | 100% every 5 years | 4,077 (66%) | 44,257 (61%) | 3.7 | 3.6 |
| 17% annually | 922 (15%) | 10,340 (14%) | 100% every 6 years | 3,744 (61%) | 40,721 (57%) | 4.1 | 3.9 |
| 14% annually | 790 (13%) | 8,863 (12%) | 100% every 7 years | 3,362 (55%) | 37,134 (52%) | 4.3 | 4.2 |
| 13% annually | 691 (11%) | 7,755 (11%) | 100% every 8 years | 3,278 (53%) | 35,485 (49%) | 4.7 | 4.6 |
| 11% annually | 615 (10%) | 6,894 (10%) | 100% every 9 years | 2,813 (46%) | 31,699 (44%) | 4.6 | 4.6 |
| 10% annually | 553 (9%) | 6,204 (9%) | 100% every 10 years | 2,777 (45%) | 30,740 (43%) | 5.0 | 5.0 |

**Women**
| Selective adherence |  |  | Sporadic adherence |  |  | Ratio sporadic / selective |  |
| --- | --- | --- | --- | --- | --- | --- | --- |
| Scheme | Prevented CRC deaths | Prevented YPLL | Scheme | Prevented CRC deaths | Prevented YPLL | Prevented CRC deaths | Prevented YPLL |
| 100% annually | 4,127 (89%) | 48,486 (87%) | 100% annually | 4,127 (89%) | 48,486 (87%) | 1.0 | 1.0 |
| 50% annually | 2,063 (44%) | 24,243 (44%) | 100% every 2 years | 3,668 (79%) | 43,350 (78%) | 1.8 | 1.8 |
| 33% annually | 1,376 (30%) | 16,162 (29%) | 100% every 3 years | 3,270 (70%) | 38,627 (69%) | 2.4 | 2.4 |
| 25% annually | 1,032 (22%) | 12,121 (22%) | 100% every 4 years | 2,937 (63%) | 34,660 (62%) | 2.8 | 2.9 |
| 20% annually | 825 (18%) | 9,697 (17%) | 100% every 5 years | 2,721 (59%) | 31,711 (57%) | 3.3 | 3.3 |
| 17% annually | 688 (15%) | 8,081 (15%) | 100% every 6 years | 2,453 (53%) | 28,814 (52%) | 3.6 | 3.6 |
| 14% annually | 590 (13%) | 6,927 (12%) | 100% every 7 years | 2,107 (45%) | 25,451 (46%) | 3.6 | 3.7 |
| 13% annually | 516 (11%) | 6,061 (11%) | 100% every 8 years | 2,118 (46%) | 24,755 (44%) | 4.1 | 4.1 |
| 11% annually | 459 (10%) | 5,387 (10%) | 100% every 9 years | 1,688 (36%) | 21,044 (38%) | 3.7 | 3.9 |
| 10% annually | 413 (9%) | 4,849 (9%) | 100% every 10 years | 1,698 (37%) | 20,622 (37%) | 4.1 | 4.3 |

Prevented CRC deaths and YPLL were calculated when compared to a scenario without screening. Numbers in brackets indicate the percentage reduction. All scenarios were simulated for a hypothetical cohort of perfectly adherent 100,000 men and women aged 50 years for up to 50 cycles of each one year. CRC: colorectal cancer. YPLL: years of potential life lost. FIT: fecal immunochemical test.

**Table 2.** Differences in Long-term Outcomes for Screening Colonoscopy at Ages 50, 60 and 70 given Varying Patterns of Adherence Yielding Identical Adherence Levels

**Men**
| Selective adherence |  |  | Sporadic adherence |  |  | Ratio sporadic / selective |  |
| --- | --- | --- | --- | --- | --- | --- | --- |
| Scheme | Prevented CRC deaths | Prevented YPLL | Scheme | Prevented CRC deaths | Prevented YPLL | Prevented CRC deaths | Prevented YPLL |
| 100% at ages 50, 60 and 70 | 5,535 (90%) | 63,582 (88%) | 100% at ages 50, 60, and 70 | 5,535 (90%) | 63,582 (88%) | 1.0 | 1.0 |
| 67% at ages 50, 60, 70 | 3,690 (60%) | 42,388 (59%) | 100% at ages 50 and 60 | 4,936 (80%) | 59,388 (83%) | 1.3 | 1.4 |
|  |  |  | 100% at ages 50 and 70 | 4,921 (80%) | 55,492 (77%) | 1.3 | 1.3 |
|  |  |  | 100% at ages 60 and 70 | 4,708 (76%) | 46,143 (64%) | 1.3 | 1.1 |
| 33% at ages 50, 60, 70 | 1,845 (30%) | 21,194 (29%) | 100% at age 50 | 3,327 (54%) | 43,784 (61%) | 1.8 | 2.1 |
|  |  |  | 100% at age 60 | 3,933 (64%) | 40,628 (56%) | 2.1 | 1.9 |
|  |  |  | 100% at age 70 | 2,989 (48%) | 22,497 (31%) | 1.6 | 1.1 |

**Women**
| Selective adherence |  |  | Sporadic adherence |  |  | Ratio sporadic / selective |  |
| --- | --- | --- | --- | --- | --- | --- | --- |
| Scheme | Prevented CRC deaths | Prevented YPLL | Scheme | Prevented CRC deaths | Prevented YPLL | Prevented CRC deaths | Prevented YPLL |
| 100% at ages 50, 60 and 70 | 4,119 (89%) | 49,613 (89%) | 100% at ages 50, 60, and 70 | 4,119 (89%) | 49,613 (89%) | 1.0 | 1.0 |
| 67% at ages 50, 60, 70 | 2,746 (59%) | 33,075 (59%) | 100% at ages 50 and 60 | 3,439 (74%) | 44,380 (80%) | 1.3 | 1.3 |
|  |  |  | 100% at ages 50 and 70 | 3,702 (80%) | 43,388 (78%) | 1.3 | 1.3 |
|  |  |  | 100% at ages 60 and 70 | 3,685 (79%) | 38,898 (70%) | 1.3 | 1.2 |
| 33% at ages 50, 60, 70 | 1,373 (30%) | 16,538 (30%) | 100% at age 50 | 2,134 (46%) | 30,601 (55%) | 1.6 | 1.9 |
|  |  |  | 100% at age 60 | 2,866 (62%) | 32,493 (58%) | 2.1 | 2.0 |
|  |  |  | 100% at age 70 | 2,638 (57%) | 22,188 (40%) | 1.9 | 1.3 |

Prevented CRC deaths and YPLL were calculated in comparison to a scenario without screening. Numbers in brackets indicate the percentage reduction. All scenarios were simulated for a hypothetical cohort of perfectly adherent 100,000 men and women aged 50 years for up to 50 cycles of each one year. CRC: colorectal cancer. YPLL: years of potential life lost

Briefly, the model simulates the natural history of CRC based on the process of precursor lesions developing into preclinical and then clinical cancer in a hypothetical population for a predefined number of years. Screening can interfere with the natural history of CRC (**Figure 1**). The model’s natural history assumptions were derived from data of the German screening colonoscopy registry, the world’s largest registry of its kind.^15–18^ CRC mortality rates were estimated using data from a German case-control study and German registry data.^10,18^ General mortality rates and average life expectancy were extracted from German population life tables.^19^

**Figure 1.**
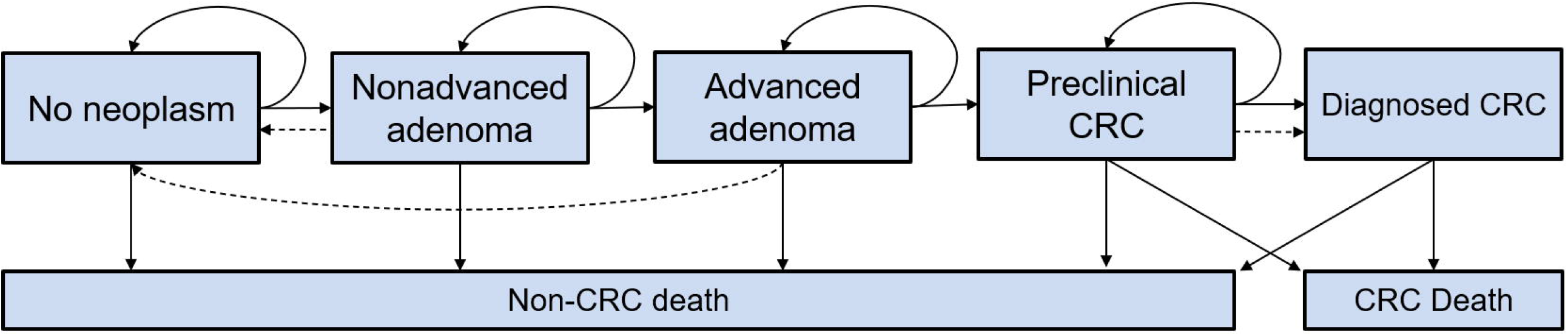
Schematic illustration of the Markov Model. Solid lines represent the progression of colorectal disease through the adenoma-carcinoma sequence in the absence of screening. Dashed lines show the movement between states because of the detection and removal of adenomas and the detection of asymptomatic CRC at screening. CRC, colorectal cancer.

The model’s external validity (its ability to reproduce real-life outcomes) was assessed by comparing model-derived cumulative incidences and prevalences to results from KolosSal, a German cohort study, registry-based estimates of CRC incidence in Germany, and outcome patterns of randomized sigmoidoscopy screening studies.^20^

The model source code, developed in the statistical software R (version 3.6.3), is available in **Supplementary Appendix 1b**.

### Simulations

#### Modelled Scenarios

In each simulated scenario, the model population consisted of previously unscreened 100,000 men and 100,000 women aged 50 at model start. Models were run for 50 years, i.e. up to age Firstly, we performed simulations assuming perfectly adhering populations for maximal offers of FIT and colonoscopy screening strategies, i.e.

1. Annual FIT screening from ages 50 to 75 years
2. Screening colonoscopy at ages 50, 60 and 70 years

Secondly, we simulated scenarios with imperfect adherence patterns for both strategies. For the purpose of this study, we defined adherence as

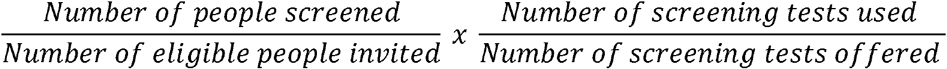

*during a given time frame*. This definition expands the definition of uptake (participation) by the European Guidelines for Quality Assurance in CRC Screening ^21^ (*number of people screened divided by the number of eligible people invited*) by a second term quantifying the frequency or intensity of (repeated) screening (*number of screening tests used divided by the number of screening tests offered*). The extension allows to assess global adherence levels over an extended period of time for situations where screening tests are offered in regular intervals, as typically the case in organized screening programs, in particular for FIT screening offers.^5^

We defined two types of distinct adherence patterns, namely *selective* and *sporadic* adherence. *Selective adherence* reflects a pattern where a selective proportion of eligible subjects adheres to screening in the recommended frequency. Hence, the second part of the equation will be 1, and the overall adherence for such a pattern equals:

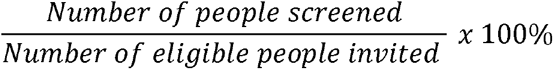

*Sporadic adherence*, on the other hand, reflects a pattern where all eligible subjects attend screening irregularly, i.e. not in the recommended frequency. Thus, for a sporadic adherence pattern, the first part of the equation will be 1, and the overall adherence equals:

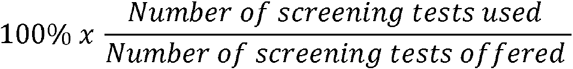

More plainly, in the case of selective adherence, the population consists of two groups, one that never uses screening and one fully adherent group. In the case of sporadic adherence, the entire population utilizes screening offers, but not at the recommended frequency. Although none of these distinct patterns is expected to occur in their ‘pure form’ in practice, the herein used clear-cut distinction allows to directly quantify the effects of different adherence patterns in terms of a screening program’s long-term efficacy.

We therefore defined, for both patterns of adherence, sets of scenarios yielding the same overall levels of adherence for the modelled time frame, i.e. 50 years. For instance, for annual FIT screening, the pair of 50% annually adherent subjects (i.e. ‘selective’ adherence) versus 100% of subjects adhering every two years (i.e. ‘sporadic’ adherence) will correspond to a level of 50% adherence, the pair of 33% annually versus 100% every three years will correspond to a level of 33% adherence, and so forth. In a similar fashion, for an offer of screening colonoscopy every ten years, the pair of 67% fully adherent subjects to screening at ages 50, 60 and 70 versus 100% adherent subjects to screening at ages 50 and 60 (or 50 and 70; or 60 and 70) will correspond to a level of 67% adherence, and the pair of 33% at ages 50, 60 and 70 versus 100% at age 50 (or 60, or 70) will correspond to a level of 33% adherence.

In all scenarios, subjects only received the next screening round if they had a negative test result in the respective previous round. Subjects with positive FITs received colonoscopy for diagnostic work-up in the same year. All subjects with either (true- or false-) positive FIT or polyps detected at screening colonoscopy received periodic surveillance colonoscopies up to age 85, which was arbitrarily chosen as upper age limit for subjects under surveillance.

#### Outcomes

For each scenario, we assessed the cumulative number of prevented deaths due to CRC after 50 years and the associated percentage reduction when compared to a scenario without screening, as well as the cumulative mortality by dividing the total number of deaths through the total simulated population. We also calculated the cumulative number of prevented years of potential life lost (YPLL) due to CRC deaths and the associated percentage reduction versus no screening. YPLL is a weighted metric taking the average remaining life expectancy at premature death into account, i.e. deaths at a younger age will be given greater weight than deaths at older age. Finally, to allow a more direct assessment of differences, we determined the ratios of prevented CRC deaths and YPLL given sporadic adherence versus the same outcome parameters given selective adherence.

### Patient and Public Involvement

Patients and the public were neither involved in the design and conduct of this study, nor in writing or editing of this document. Research at the German Cancer Research Center (DKFZ) is generally informed by a Patient Advisory Committee.

## Results

Table 1 and **Table 2** show differences in simulated outcomes after 50 years given varying patterns of adherence for screening with FIT and colonoscopy, respectively. Trajectories of cumulative mortality and cumulative YPLL at selected levels of adherence are shown in **Figure 2** for annual FIT screening, and in **Figures 3 and 4** for screening colonoscopy at ages 50, 60 and 70.

Assuming a perfectly adhering population, both annual FIT screening and screening colonoscopy at ten-year intervals resulted in pronounced reductions of CRC deaths (89-90%) and YPLL due to CRC (86-90%) as compared to no screening in both sexes.

**Figure 2.**
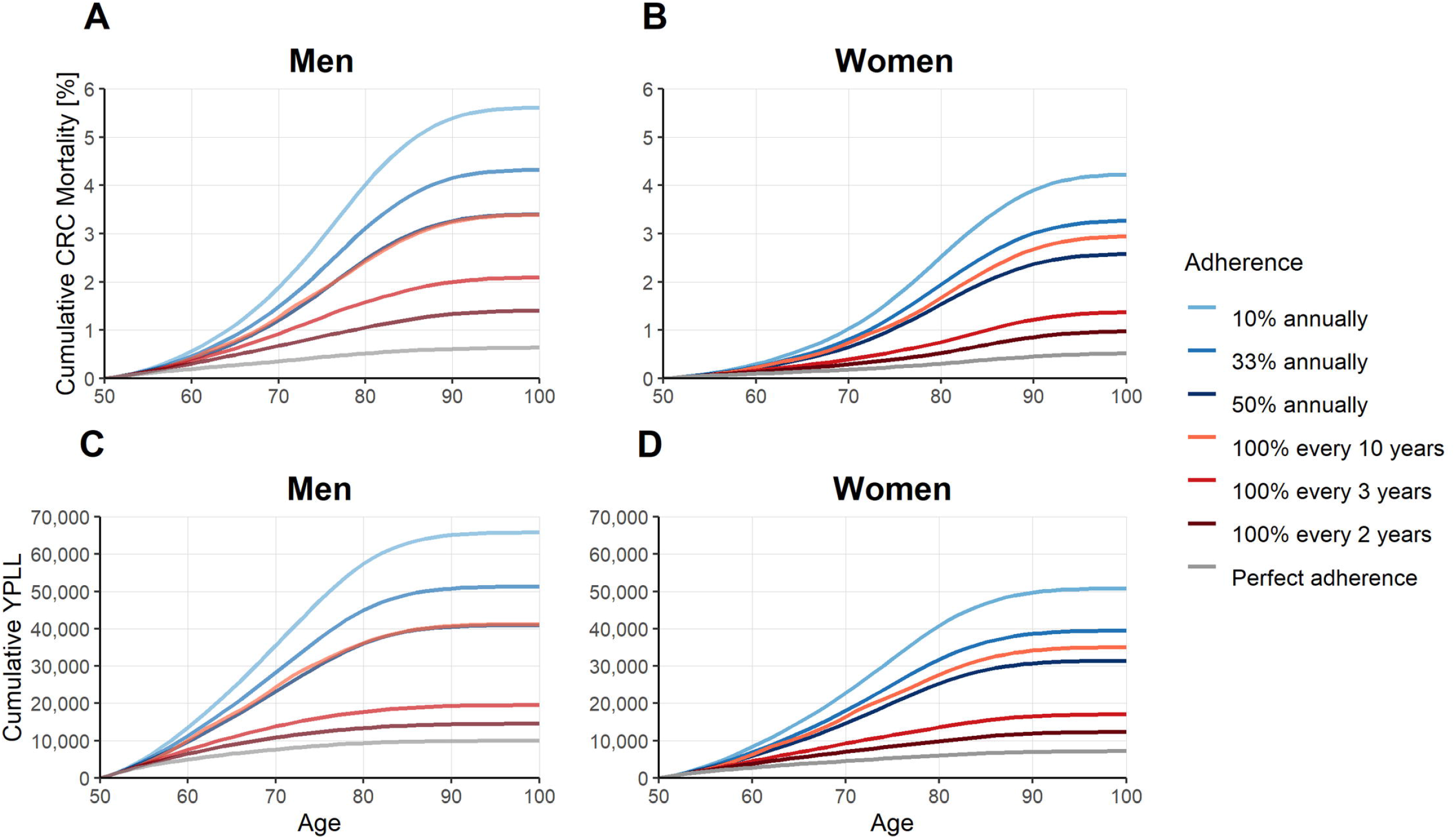
Cumulative Mortality (A, B) and Cumulative YPLL due to CRC Deaths (C, D) for Annual FIT Screening Given Varying Patterns of Adherence at Selected Adherence Levels (10%, 33%, 50%), Stratified by Sex. Blue shades: Selective adherence (only a part of the population utilizes screening offers, but in the recommended frequency) Red shades: Sporadic adherence (the entire population utilizes screening offers, but not in the recommended frequency) CRC, colorectal cancer; FIT, fecal immunochemical test; YPLL, years of potential life lost

**Figure 3.**
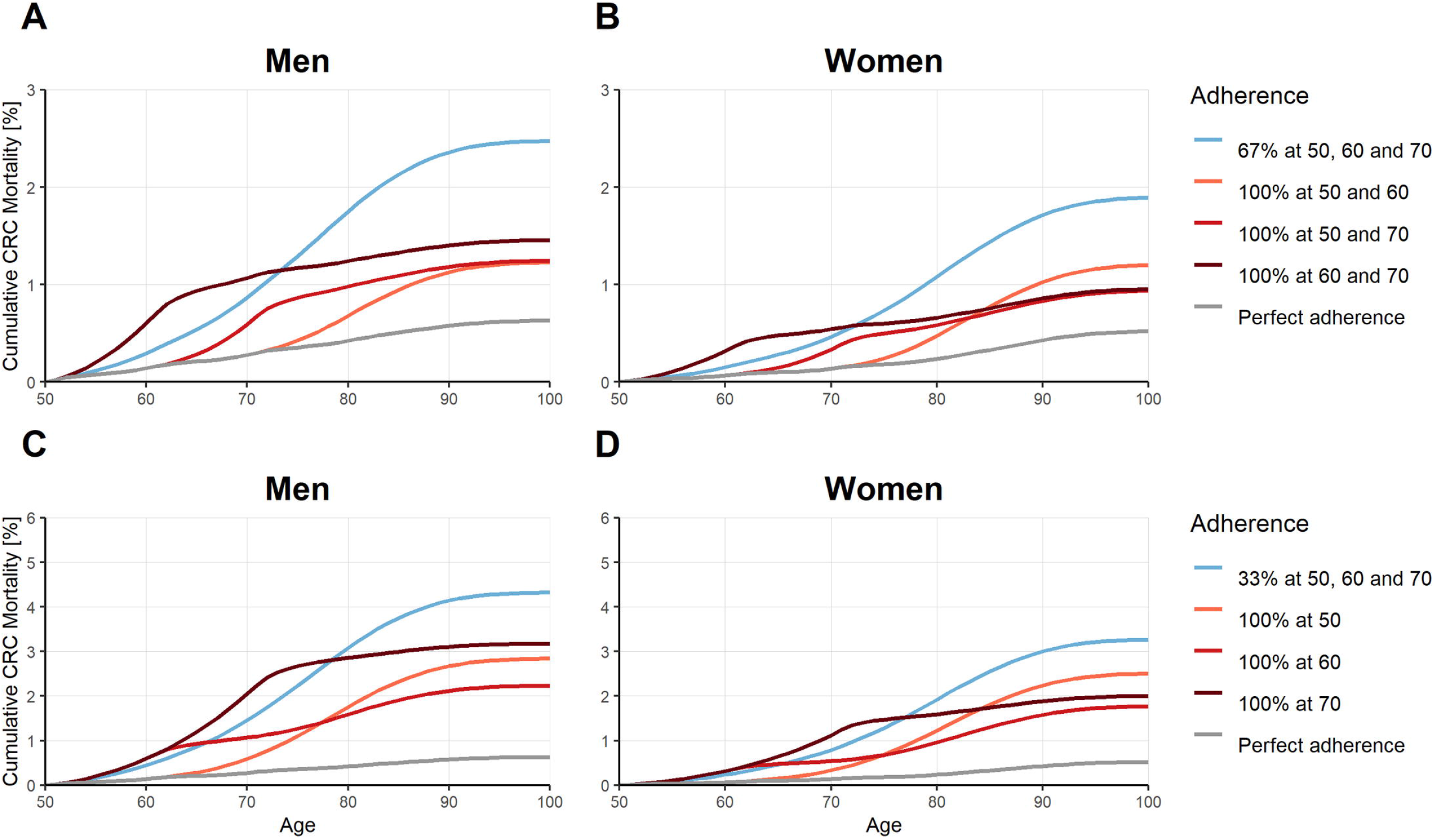
Cumulative Mortality for Screening Colonoscopy at Ages 50, 60 and 70 Given Varying Patterns of Adherence at Selected Adherence Levels (67%, A and B; and 33%, C and D), Stratified by Sex. Blue: Selective adherence (only a part of the population utilizes screening offers, but in the recommended frequency) Red shades: Sporadic adherence (the entire population utilizes screening offers, but not in the recommended frequency) CRC, colorectal cancer

**Figure 4.**
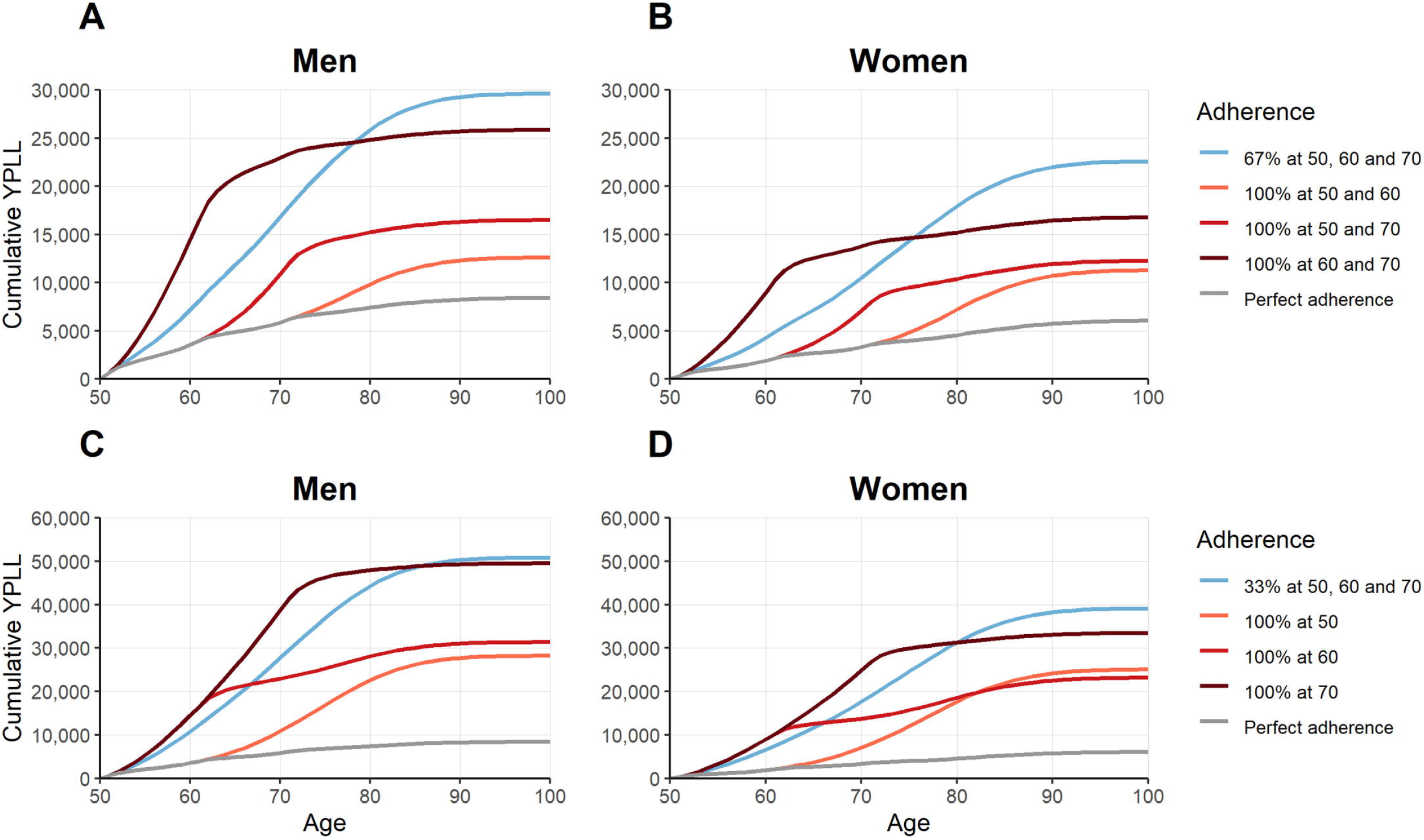
Cumulative YPLL for Screening Colonoscopy at Ages 50, 60 and 70 Given Varying Patterns of Adherence at Selected Adherence Levels (67%, A and B; and 33%, C and D), Stratified by Sex. Blue: Selective adherence (only a part of the population utilizes screening offers, but in the recommended frequency) Red shades: Sporadic adherence (the entire population utilizes screening offers, but not in the recommended frequency) CRC, colorectal cancer; YPLL, years of potential life lost

### Annual FIT Screening

Different patterns of adherence, while at the same overall adherence levels, were associated with considerable differences in screening efficacy. For annual FIT screening at simulated adherence levels of up to 50%, ratios of prevented CRC deaths assuming sporadic versus selective patterns ranged from 1.9-5.0 for men and from 1.8-4.1 for women, i.e., at the same overall level of adherence, in men up to 5-times and in women up to 4 times more CRC deaths were prevented when the simulated population followed a sporadic rather than a selective adherence pattern. Differences were similar for YPLL (ratios sporadic versus selective, 1.9-5.0 in men, 1.8-4.3 in women). Overall, ratios calculated for outcomes assuming sporadic versus selective adherence tended to be less pronounced for comparably high (e.g. 50%) and more pronounced for low (e.g. 10%) levels of adherence and tended to be higher in men as compared to women.

### Screening Colonoscopy at Ten-Year Intervals

Assuming adherence levels of 33% and 67% for screening colonoscopy at ten-year intervals revealed a similar trend as observed for annual FIT screening, i.e. the sporadic scheme yielded overall higher levels of efficacy than the selective scheme. Ratios of prevented CRC deaths for sporadic versus selective patterns ranged from 1.3-2.1 for both sexes, and ratios for prevented YPLL ranged from 1.1-2.1 in men and 1.2-2.0 in women. They tended to be higher for men in case of sporadic adherence at younger ages and higher for women in case of sporadic adherence at older ages.

## Discussion

This simulation study provided estimates on the efficacy of colorectal cancer screening strategies based on varying patterns of the eligible population’s screening behavior. Assuming distinct adherence patterns, defined as *selective adherence* (where only a part of the population utilizes screening offers, but in the recommended frequency) and *sporadic adherence* (where the entire population utilizes screening offers, but not in the recommended frequency), we found that different patterns of adherence, while at the same overall adherence level, were associated with considerable differences in long-term efficacy. Our findings suggest that substantially more CRC deaths and YPLL could be prevented when the population followed a sporadic instead of a selective adherence pattern. For annual FIT screening (at uptake levels of 10-50%) and for screening colonoscopy at ten-year intervals (at uptake levels of 33% and 67%) adopting a population-wide sporadic adherence pattern was estimated to be up to 4-5 times and up to 2 times more effective than a selective pattern, respectively.

### Findings in Context

Population-based screening for colorectal cancer involves a multitude of complex programmatic issues, reflected in differences in the design of screening programs around the world. Programs vary with respect to the eligible population, targeted age groups, implementation approach, objectives as well as offered screening tests and intervals.^5^ Eventually, however, the effectiveness of any program will be driven by two main components, firstly the efficacy of the offered screening tests when they are actually used, and secondly their uptake by the targeted population.

So far, it remains unclear which screening strategy is the most effective to offer. No direct comparisons of the performance of alternative strategies have been completed, and long-term outcomes of four head-to-head studies underway are not expected before the late 2020s.^22–25^ Evidence from modelling studies based on perfect adherence, an assumption which reflects the point of view of an individual subject, suggest that screening with colonoscopy at ten year intervals may be the most efficacious strategy, but other strategies, including annual FIT, do not fall much behind in terms of the reduction of the risk of dying from CRC.^26^

From a public health perspective, achieving high levels of population adherence is an additional and particularly important component to be considered. Health authorities may decide to introduce one screening strategy or another, e.g. annual FIT or ten-yearly colonoscopy, as assumed in our study, but the eligible population’s uptake can at best be influenced indirectly. The necessary understanding of the mechanisms of adherence to colorectal cancer screening offers is constantly evolving. Broadly, characteristics of eligible population and program design are known to impact the population uptake. For instance, higher participation rates have been reported among women versus men and in those aged older than 60 years versus younger age groups.^8^ An association was also reported for lifestyle factors and cultural background.^11^

A recent study found that European countries with nationwide coverage of organized screening programs offering only fecal testing or a fecal testing and colonoscopy as alternatives had the highest levels of population adherence.^8^ Higher uptake of FIT versus colonoscopy was also seen in the first round of the randomized COLONPREV study, comparing one-time colonoscopy versus biennial FIT.^22^ Uptake in subsequent screening rounds tends to be high among those adherent to a first invitation.^27,28^ Several observational and randomized studies investigating approaches to increase uptake have consistently shown that personal invitations with directly mailed FITs lead to higher utilization rates.^29,30^ Patient navigation and education were also found to have a significant effect.^31^

Differential patterns of screening behavior through several screening rounds have been described previously.^32,33^ These include consistent screening attendees and consistent non-responders (reflected in the ideal-typical selective adherence pattern in our study), as well as intermittent attendees with late entry, drop out, or intermittent participation (reflected in the ideal-typical sporadic adherence pattern). Estimates on the distribution of these behavioral patterns within a specific population are naturally difficult, given the limited evidence and large heterogeneity of population characteristics. In the few available studies, approximately 40-50% of studied subjects were consistent screeners and 20-30% consistent non-responders, with the remainder following intermittent participation patterns.^27,28,32–34^

While it has been acknowledged that differences in adherence patterns need to be taken into account when interpreting trial results,^26^ the evidence on the actual impact on long-term outcomes is scant. A retrospective cohort study found that not being up to date in screening increased the risk for CRC death nearly 3-fold.^14^ A previous modeling study suggested that approximately 60% of US CRC deaths are attributable to nonuse of screening.^35^ To our knowledge, no previous study has assessed differential longitudinal adherence patterns in terms of their impact on long-term outcomes.

Our study adds to the literature that, from a societal perspective, large proportions of the population making sporadic use of screening offers will be substantially more beneficial to achieve sustained reductions of CRC mortality and YPLL due to CRC deaths than small proportions of the population utilizing screening offers at the recommended frequency. Notably, the intention of our study was to illustrate differences in efficacy driven by distinct adherence patterns within exemplary FIT or colonoscopy-based strategies, and not to compare outcomes of these strategies to each other. For this reason, assessed levels of imperfect adherence were chosen for their convenience of illustration, and therefore vary between the strategies (10-50% for FIT; 33% and 67% for colonoscopy). The overall adherence level used for the specific analysis should be kept in mind when interpreting the results; for instance, the 4-5 times higher reductions of CRC deaths and YPLL achieved under a sporadic vs. a selective FIT adherence scheme were found at overall adherence levels of 10-20%, which was not assessed for screening colonoscopy.

### Implications for Colorectal Cancer Screening

Clearly, optimal protection from CRC can only be achieved by perfect adherence. However, even in countries with well-organized screening programs, such as the Netherlands, where up to 60% of the eligible population regularly attend screening,^36,37^ full adherence remains far out of reach. Therefore, in situations of limited resources, our findings should encourage health authorities to concentrate efforts on promoting broader reach of screening in the eligible population, paying particular attention to efforts to better target and reach notoriously more difficult-to-reach population groups, such as less educated or otherwise socially disadvantaged groups.^38^

Finally, our study points to the need of a broader reflection on the long-term effects of adherence mechanisms. European guidelines consider a minimum uptake of 45% as acceptable but recommend aiming for at least 65%.^21^ In the US, the National Colorectal Cancer Roundtable had agreed on a goal of 80% screened.^39^ Suchlike aspirational targets typically refer to the number of people screened as a proportion of all people who are invited to attend a specific screening offer within population-based screening programs. Accordingly, screening adherence is usually quantified as the proportion of the population up to date with screening, e.g., that has had a FIT within the past 1 or 2 years, or a colonoscopy within the past 10 years.^7–9^ Our results suggest expanding such adherence metrics by additional indicators taking adherence patterns over multiple rounds of screening into account whenever the data allow to do so. Ideally, such metrics should inform on ‘effective’ adherence, e.g. by reporting the proportion of subjects who ever used a screening test, or the re-screening adherence when compared to one or several previous rounds of screening. Where reporting of longitudinal uptake rates is incomplete or lacking, our study may also contribute to understand differences in effectiveness of comparably designed screening programs despite seemingly comparable overall uptake levels.

### Strengths and Limitations

Specific strengths and limitations of the model per se have been described previously.^10,20,40^ Briefly, a major strength is the use of input parameters derived specifically from the German general population using the world’s largest screening colonoscopy registry. Furthermore, the model was subjected to a thorough assessment of its external validity, and was found to adequately predict colorectal neoplasm prevalences and incidences in a German population, with estimated patterns of the effect of screening colonoscopy resembling those seen in registry data and real-world studies.^20^ Major limitations concern model simplifying assumptions and uncertainties related to input parameters where evidence was limited, for instance with regard to transition rates for age groups 50-54 years and 80+ years, true screening test performance characteristics in Germany and potential differences between sexes in this respect. In addition, the chosen approach of a Markov-type simulation model is very well suited to quantify average effects on the level of populations, but less so for individuals potentially at substantially lower or higher risk of CRC. Key input parameters of our model were derived from the general German population, which therefore implies that inference from our findings should be restricted to the screening eligible population at average risk, i.e. subjects without personal history of CRC or adenomas, no inflammatory bowel disease, no hereditary CRC syndrome, and no family history of CRC.

## Data Availability

Availability of data, code and material:
All analyses relevant to the study are included in the article or uploaded as supplementary information. The model source code is uploaded as supplementary information.

## Conclusion

In summary, long-term outcomes of colorectal cancer screening programs may vary substantially due to underlying longitudinal patterns of the population’s screening behavior. In an average-risk screening population, sporadic use of screening offers by a large part of the eligible population is likely substantially more beneficial to achieve sustained reductions of the CRC burden than intensive use of screening offers by a small part of the population. Adherence statistics should be refined to better reflect ‘effective adherence’.

## References

1. Hewitson P, Glasziou P, Watson E, et al. Cochrane systematic review of colorectal cancer screening using the fecal occult blood test (hemoccult): an update. Am J Gastroenterol 2008;103:1541–9.

2. Brenner H, Stock C, Hoffmeister M. Effect of screening sigmoidoscopy and screening colonoscopy on colorectal cancer incidence and mortality: systematic review and meta-analysis of randomised controlled trials and observational studies. BMJ 2014;348:g2467.

3. Senore C, Hassan C, Regge D, et al. Cost-effectiveness of colorectal cancer screening programmes using sigmoidoscopy and immunochemical faecal occult blood test. J Med Screen 2019;26:76–83.

4. Ran T, Cheng CY, Misselwitz B, et al. Cost-Effectiveness of Colorectal Cancer Screening Strategies-A Systematic Review. Clin Gastroenterol Hepatol 2019;17:1969-1981.e15.

5. Schreuders EH, Ruco A, Rabeneck L, et al. Colorectal cancer screening: a global overview of existing programmes. Gut 2015;64:1637–49.

6. Bénard F, Barkun AN, Martel M, et al. Systematic review of colorectal cancer screening guidelines for average-risk adults: Summarizing the current global recommendations. World J Gastroenterol 2018;24:124–138.

7. Levin TR, Corley DA, Jensen CD, et al. Effects of Organized Colorectal Cancer Screening on Cancer Incidence and Mortality in a Large Community-Based Population. Gastroenterology 2018;155:1383-1391.e5.

8. Cardoso R, Guo F, Heisser T, et al. Utilisation of Colorectal Cancer Screening Tests in European Countries by Type of Screening Offer: Results from the European Health Interview Survey. Cancers 2020;12:1409.

9. Joseph DA. Vital Signs: Colorectal Cancer Screening Test Use — United States, 2018. MMWR Morb Mortal Wkly Rep 2020;69. Available at: https://www.cdc.gov/mmwr/volumes/69/wr/mm6910a1.htm [Accessed June 17, 2020].

10. Chen C, Stock C, Hoffmeister M, et al. Optimal age for screening colonoscopy: a modeling study. Gastrointest Endosc 2019;89:1017-1025.e12.

11. Klabunde C, Blom J, Bulliard J-L, et al. Participation rates for organized colorectal cancer screening programmes: an international comparison. J Med Screen 2015;22:119–126.

12. Cardoso R, Niedermaier T, Chen C, et al. Colonoscopy and Sigmoidoscopy Use among the Average-Risk Population for Colorectal Cancer: A Systematic Review and Trend Analysis. Cancer Prev Res (Phila) 2019;12:617–630.

13. Senore C, Basu P, Anttila A, et al. Performance of colorectal cancer screening in the European Union Member States: data from the second European screening report. Gut 2019;68:1232–1244.

14. Doubeni CA, Fedewa SA, Levin TR, et al. Modifiable Failures in the Colorectal Cancer Screening Process and Their Association With Risk of Death. Gastroenterology 2019;156:63-74.e6.

15. Brenner H, Altenhofen L, Stock C, et al. Prevention, early detection, and overdiagnosis of colorectal cancer within 10 years of screening colonoscopy in Germany. Clin Gastroenterol Hepatol 2015;13:717–23.

16. Brenner H, Altenhofen L, Stock C, et al. Expected long-term impact of the German screening colonoscopy programme on colorectal cancer prevention: Analyses based on 4,407,971 screening colonoscopies. Eur J Cancer 2015;51:1346–1353.

17. Brenner H, Kretschmann J, Stock C, et al. Expected long-term impact of screening endoscopy on colorectal cancer incidence: A modelling study. Oncotarget 2016;7:48168–48179.

18. Chen C, Stock C, Hoffmeister M, et al. How long does it take until the effects of endoscopic screening on colorectal cancer mortality are fully disclosed?: a Markov model study. Int J Cancer 2018;143:2718–2724.

19. Rößger F. General Life Table 2010/2012. Methodological Description and Results (Allgemeine Sterbetafel 2010/2012. Methodische Erläuterungen und Ergebnisse). Wiesbaden: Federal Office of Statistics (Statistisches Bundesamt); 2015. Available at: www-genesis.destatis.de.

20. Heisser T, Hoffmeister M, Brenner H. Effects of Screening for Colorectal Cancer: Development and Validation of a Multistate Markov Model. medRxiv 2020:2020.04.17.20069484.

21. Moss S, Ancelle-Park R, Brenner H, et al. European guidelines for quality assurance in colorectal cancer screening and diagnosis. First Edition--Evaluation and interpretation of screening outcomes. Endoscopy 2012;44 Suppl 3:SE49–64.

22. Quintero E, Castells A, Bujanda L, et al. Colonoscopy versus Fecal Immunochemical Testing in Colorectal-Cancer Screening. N Engl J Med 2012;366:697–706.

23. Bretthauer M, Kaminski MF, Loberg M, et al. Population-Based Colonoscopy Screening for Colorectal Cancer: A Randomized Clinical Trial. JAMA Intern Med 2016;176:894–902.

24. Dominitz JA, Robertson DJ, Ahnen DJ, et al. Colonoscopy vs. Fecal Immunochemical Test in Reducing Mortality From Colorectal Cancer (CONFIRM): Rationale for Study Design. Am J Gastroenterol 2017;112:1736–1746.

25. Hultcrantz R. Colonoscopy and FIT as Colorectal Cancer Screening Test in the Average Risk Population. ClinicalTrials.gov Identifier: NCT02078804. 2020. Available at: www.clinicaltrials.gov [Accessed June 17, 2020].

26. Ladabaum U, Dominitz JA, Kahi C, et al. Strategies for Colorectal Cancer Screening. Gastroenterology 2020;158:418–432.

27. Jensen CD, Corley DA, Quinn VP, et al. Fecal Immunochemical Test Program Performance Over 4 Rounds of Annual Screening: A Retrospective Cohort Study. Ann Intern Med 2016;164:456–463.

28. Zorzi M, Hassan C, Capodaglio G, et al. Long-term performance of colorectal cancerscreening programmes based on the faecal immunochemical test. Gut 2018;67:2124–2130.

29. Mehta SJ, Pepe RS, Gabler NB, et al. Effect of Financial Incentives on Patient Use of Mailed Colorectal Cancer Screening Tests: A Randomized Clinical Trial. JAMA Netw Open 2019;2:e191156–e191156.

30. Gruner LF, Hoffmeister M, Ludwig L, et al. Effect of Various Invitation Schemes on the Use of Fecal Immunochemical Tests for Colorectal Cancer Screening: Protocol for a Randomized Controlled Trial. JMIR Res Protoc 2020;9. Available at: https://www.ncbi.nlm.nih.gov/pmc/articles/PMC7165303/ [Accessed June 7, 2020].

31. Dougherty MK, Brenner AT, Crockett SD, et al. Evaluation of Interventions Intended to Increase Colorectal Cancer Screening Rates in the United States. JAMA Intern Med 2018;178:1645–1658.

32. Duncan A, Turnbull D, Wilson C, et al. Behavioural and demographic predictors of adherence to three consecutive faecal occult blood test screening opportunities: a population study. BMC Public Health 2014;14:238.

33. Lo SH, Halloran S, Snowball J, et al. Colorectal cancer screening uptake over three biennial invitation rounds in the English bowel cancer screening programme. Gut 2015;64:282–291.

34. Vlugt M van der, Grobbee EJ, Bossuyt PM, et al. Adherence to colorectal cancer screening: four rounds of faecal immunochemical test-based screening. Br J Cancer 2017;116:44–49.

35. Meester RGS, Doubeni CA, Lansdorp-Vogelaar I, et al. Colorectal cancer deaths attributable to nonuse of screening in the United States. Annals of Epidemiology 2015;25:208-213.e1.

36. Roon AHC van, Hol L, Wilschut JA, et al. Advance notification letters increase adherence in colorectal cancer screening: A population-based randomized trial. Preventive Medicine 2011;52:448–451.

37. Toes-Zoutendijk E, Leerdam ME van, Dekker E, et al. Real-Time Monitoring of Results During First Year of Dutch Colorectal Cancer Screening Program and Optimization by Altering Fecal Immunochemical Test Cut-Off Levels. Gastroenterology 2017;152:767-775.e2.

38. Chen C, Hoffmeister M, Brenner H. The toll of not screening for colorectal cancer. Expert Review of Gastroenterology & Hepatology 2017;11:1–3.

39. National Colorectal Cancer Roundtable. Working toward the shared goal of 80% screened for colorectal cancer by 2018. Available at: http://nccrt.org/tools/80-percent-by-2018/.

40. Heisser T, Weigl K, Hoffmeister M, et al. Age-specific sequence of colorectal cancer screening options in Germany: A model-based critical evaluation. PLOS Medicine 2020;17:e1003194.

