## Supplementary Appendix 1 for "Strongly Divergent Impact of Adherence Patterns on Efficacy of Colorectal Cancer Screening: The Need to Refine Adherence Statistics"

### **SUPPLEMENTARY MATERIAL**

**Strongly Divergent Impact of Adherence Patterns**

**on Efficacy of Colorectal Cancer Screening:**

**The Need to Refine Adherence Statistics**

Efficacy of Adherence Patterns

Thomas Heisser, Rafael Cardoso, Feng Guo, Tobias Moellers, Michael Hoffmeister, Hermann Brenner

**Supplementary Appendix 1a: Model Documentation.**

**Supplementary Appendix 1b: R Code.**

#### **Supplementary Appendix 1a. Model Documentation**

##### **Conceptual model structure**

Our multistate Markov model simulates the natural history of CRC based on the process of precursor lesions (non-advanced and advanced adenomas) developing into preclinical (asymptomatic) and then clinical (symptomatic) cancer. The simulation is performed on a hypothetical previously unscreened German population, with the number of simulated subjects and their corresponding baseline age (minimum 50 years) being variables to be chosen prior to model start. The model can principally be used for simulating any population, provided updated or appropriately adjusted input parameters.

At start of the simulation, certain proportions of no neoplasm, non-advanced adenoma, advanced adenoma and preclinical CRC are assigned to the hypothetical population. The simulation runs up to a predefined number of cycles of each one year. Each year, people at each state have a certain probability (transition rate) to progress to the next state. Subjects with CRC may die from the disease, and at each state people may experience non-CRC death, reflecting the general background mortality from other causes.

Screening can alter the progression between states. People with adenoma will be moved backward to the state of no neoplasm, assuming removal of their adenoma at colonoscopy (for screening or diagnostic workup, e.g. after a positive fecal test). Subjects will then continue to have the probabilities to progress to the next states as those without findings at screening. We assume that, although these people are under a higher risk of developing adenomas or cancers than the general population,^1^ the excess risk will be effectively compensated through the protection provided by surveillance colonoscopies.^2,3^ Preclinical CRC detected at screening will be moved forward to the state of diagnosed cancer.

After each cycle where a screening test was applied, the model is able to differentiate the simulated population into a ‘screening negative’ and a ‘screening positive’ group, which allows for modelling different trajectories depending on the screening outcome.

##### **Model parameters**

**Starting prevalences and transition rates**

An overview of key model parameters is given in **Appendix Table 1**.

*Data source*

The data basis of our analyses on model starting prevalences and transition rates was the nationwide screening colonoscopy registry run by the Central Research Institute of Ambulatory Health Care in Germany. The registry, which was built up along with the introduction of the screening colonoscopy offer in the year 2002, is a repository of all screening colonoscopies conducted in Germany. Reporting is virtually complete, as it is a prerequisite for physicians’ reimbursement by the health insurance funds. The registry includes only primary screening examinations (i.e., colonoscopies conducted for surveillance, work-up of symptoms or other screening tests are not included). Items reported include, besides basic sociodemographic variables, findings at colonoscopy, including number, size and histological characteristics of polyps. In case of multiple neoplasms, only the most advanced one (non-advanced adenoma, advanced adenoma, or cancer) is recorded. Advanced adenomas are defined as at least 1 adenoma ≥ 1 cm or at least 1 adenoma with villous components or high-grade dysplasia.

Noteworthy, the reporting for the screening colonoscopy registry does not differentiate by the class of lesion. Thus, the herein used term ‘adenoma’ refers to conventional or serrated adenomas (polyps) alike. While we preferred to refer to our model as being based on the adenoma-carcinoma pathway in previous publications ^4–8^ for the sake of simplicity and comprehensibility (as the grand majority of CRCs develops through this well-established pathway of cancer development ^9,10^), in fact the model’s defining parameters were derived using polyp/adenoma prevalences as detected and reported at screening colonoscopy, regardless of their underlying mechanism or pathway of development. Therefore, it will be more precise to refer to the model as being based on the ‘natural history of CRC’, without restrictions on underlying CRC development pathways.

*Starting prevalence*

The proportions of no neoplasm, non-advanced adenoma, advanced adenoma and preclinical CRC at the beginning of simulation were calculated based on the data from 344,658 participants of the German screening colonoscopy program who had their first screening colonoscopy during 2003–2012 at the age of 55 years.^6^ To take into account that a certain proportion of neoplasms needs to be assumed to have been missed at colonoscopy screening, in particular for serrated or flat polyps,^11,12^ we re-calculated the previously reported prevalences,^6^ assuming representative miss rates of 25% for non-advanced adenomas and 5% for advanced neoplasms (advanced adenomas and preclinical cancers).

This was used as the best estimate for simulations starting with a 50-year-old population, which seems reasonable as selected regional programs which offer screening colonoscopy from age 50 on found similar prevalences of adenomas in age groups 50-54 and 55-59 ^13^.

*Transition rates*

Transition rates between states were also estimated based on data from the nationwide screening colonoscopy registry. Same as for the starting prevalences, we re-calculated previously reported transition rates ^14–16^ to adjust for representative colonoscopy miss rates ^11,12^. Age- and sex-specific annual transition rates between the states were estimated for age groups from 55-79 years in steps of 5 years. Estimates for age 50-54 and ≥ 80 (or ≥ 85) were assumed to be the same as those for age group 55-59 and 75-79 (or 80-84), respectively.

Confidence intervals for both starting prevalences and transition rates were derived by bootstrap analysis with resampling within sex- and age-specific subgroups. Ninety-five percent confidence intervals were determined as the 2.5th and 97.5th percentile of transition rate estimates obtained in 1,000 runs.

**Mortality rates**

Mortality rates for patients whose cancer was detected by screening or by symptoms were estimated in previous analyses ^7,8^. We combined data on the proportion of screening-detected cases among all CRC cases in Germany during 2003-2012 in people aged 55-79 years^4,17^ with the overall CRC-specific mortality rates by year after diagnosis in Germany in 2011-2012.^17^ We then used hazard ratios for patients detected by screening versus symptoms as obtained from a German population-based case-control study on CRC screening with long-term mortality follow-up of CRC patients ^7,18^ to estimate CRC-specific mortality rates by mode of detection (**Appendix Table 2**). Sex- and age-specific general mortality rates and average life expectancy of the population were extracted from German population life tables 2010/2012 (**Appendix Table 3)** ^19^.

**Diagnostic performance**

*Colonoscopy*

For colonoscopy, we assumed representative sensitivities (the proportion of detected cases among all subjects with the disease) of 75% and 95% for non-advanced adenomas and advanced neoplasms, respectively, based on evidence on the polyp and adenoma miss rates determined by tandem colonoscopy ^11,12^, with perfect specificity (the proportion of healthy people classified as such among all subjects without the disease) (100%). As our model follows a population-based approach which implies assigning global parameters to all subjects, this already incorporates differences in miss rates found according to polyp class.

*FIT*

The sensitivity and specificity of FIT screening were adjusted for a positivity rate of 10% as reported by an analysis by Gies and colleagues who showed that the positivity rate is a very good proxy indicator for the diagnostic performance ^20,21^. We chose to use the sensitivity and specificity parameters (**Appendix Table 1C**) as adjusted for a positive rate of 10% as this was the overall positivity rate of FITs used in Germany in 2018 ^22^ and assumed an absolute difference of 5% in sensitivities (higher in men) and specificities (higher in women) based on evidence suggesting varying performance according to sex ^23–26^. For the base case scenario, due to the lack of evidence suggesting otherwise and in line with previous models on FIT screening ^27,28^, conditional independence between repeated rounds of FIT testing was assumed.

##### **References**

1.	Cottet V, Jooste V, Fournel I, et al. Long-term risk of colorectal cancer after adenoma removal: a population-based cohort study. Gut 2012;61:1180–6.

2.	Lieberman DA, Rex DK, Winawer SJ, et al. Guidelines for colonoscopy surveillance after screening and polypectomy: a consensus update by the US Multi-Society Task Force on Colorectal Cancer. Gastroenterology 2012;143:844–857.

3.	Hassan C, Quintero E, Dumonceau JM, et al. Post-polypectomy colonoscopy surveillance: European Society of Gastrointestinal Endoscopy (ESGE) Guideline. Endoscopy 2013;45:842–51.

4.	Brenner H, Altenhofen L, Stock C, et al. Prevention, early detection, and overdiagnosis of colorectal cancer within 10 years of screening colonoscopy in Germany. Clin Gastroenterol Hepatol 2015;13:717–23.

5.	Brenner H, Altenhofen L, Stock C, et al. Expected long-term impact of the German screening colonoscopy programme on colorectal cancer prevention: Analyses based on 4,407,971 screening colonoscopies. Eur J Cancer 2015;51:1346–1353.

6.	Brenner H, Kretschmann J, Stock C, et al. Expected long-term impact of screening endoscopy on colorectal cancer incidence: A modelling study. Oncotarget 2016;7:48168–48179.

7.	Chen C, Stock C, Hoffmeister M, et al. How long does it take until the effects of endoscopic screening on colorectal cancer mortality are fully disclosed?: a Markov model study. Int J Cancer 2018;143:2718–2724.

8.	Chen C, Stock C, Hoffmeister M, et al. Optimal age for screening colonoscopy: a modeling study. Gastrointest Endosc 2019;89:1017-1025.e12.

9.	Dekker E, Tanis PJ, Vleugels JLA, et al. Colorectal cancer. Lancet 2019;394:1467–1480.

10.	Winawer SJ. Natural history of colorectal cancer. Am J Med 1999;106:3–6.

11.	Rijn JC van, Reitsma JB, Stoker J, et al. Polyp miss rate determined by tandem colonoscopy: a systematic review. Am J Gastroenterol 2006;101:343–50.

12.	Zhao S, Wang S, Pan P, et al. Magnitude, Risk Factors, and Factors Associated With Adenoma Miss Rate of Tandem Colonoscopy: A Systematic Review and Meta-analysis. Gastroenterology 2019;156:1661-1674.e11.

13.	Brenner H, Zwink N, Ludwig L, et al. Should Screening Colonoscopy Be Offered From Age 50? Dtsch Arztebl Int 2017;114:94–100.

14.	Brenner H, Altenhofen L, Katalinic A, et al. Sojourn Time of Preclinical Colorectal Cancer by Sex and Age: Estimates From the German National Screening Colonoscopy Database. Am J Epidemiol 2011;174:1140–1146.

15.	Brenner H, Altenhofen L, Stock C, et al. Natural history of colorectal adenomas: birth cohort analysis among 3.6 million participants of screening colonoscopy. Cancer Epidemiol Biomarkers Prev 2013;22:1043–51.

16.	Brenner H, Altenhofen L, Stock C, et al. Incidence of colorectal adenomas: birth cohort analysis among 4.3 million participants of screening colonoscopy. Cancer Epidemiol Biomarkers Prev 2014;23:1920–7.

17.	Anon. Cancer Statistics for Germany. Interactive Database of the German Centre for Cancer Registry Data (ZfKD). 2019. Available at: www.krebsdaten.de [Accessed November 20, 2019].

18.	Weigl K, Jansen L, Chang-Claude J, et al. Family history and the risk of colorectal cancer: The importance of patients’ history of colonoscopy. Int J Cancer 2016;139:2213–20.

19.	Rößger F. *General Life Table 2010/2012. Methodological Description and Results (Allgemeine Sterbetafel 2010/2012. Methodische Erläuterungen und Ergebnisse).* Wiesbaden: Federal Office of Statistics (Statistisches Bundesamt); 2015. Available at: www-genesis.destatis.de.

20.	Gies A, Bhardwaj M, Stock C, et al. Quantitative fecal immunochemical tests for colorectal cancer screening. Int J Cancer 2018;143:234–244.

21.	Gies A, Cuk K, Schrotz-King P, et al. Direct Comparison of Diagnostic Performance of 9 Quantitative Fecal Immunochemical Tests for Colorectal Cancer Screening. Gastroenterology 2018;154:93–104.

22.	Federal Joint Committee (Gemeinsamer Bundesausschuss). *The iFOBT in Colorectal Cancer Screening: Results from Medicinal Laboratories for the Year 2018. (Der	iFOBT	im	Darmkrebs-Screening: Ergebnisse	der	medizinischen	Laboratorien für das Jahr 2018)*. Berlin; 2018. Available at: www.g-ba.de/downloads/17-98-4777/2019-03-25_G-BA_iFOBT_Quartalsbericht_2018_.pdf.

23.	Brenner H, Haug U, Hundt S. Sex differences in performance of fecal occult blood testing. Am J Gastroenterol 2010;105:2457–2464.

24.	Khalid-de Bakker CAJ, Jonkers DMAE, Sanduleanu S, et al. Test performance of immunologic fecal occult blood testing and sigmoidoscopy compared with primary colonoscopy screening for colorectal advanced adenomas. Cancer Prev Res (Phila) 2011;4:1563–1571.

25.	Grobbee EJ, Wieten E, Hansen BE, et al. Fecal immunochemical test-based colorectal cancer screening: The gender dilemma. United European Gastroenterol J 2017;5:448–454.

26.	Brenner H, Qian J, Werner S. Variation of diagnostic performance of fecal immunochemical testing for hemoglobin by sex and age: results from a large screening cohort. Clin Epidemiol 2018;10:381–389.

27.	Knudsen AB, Zauber AG, Rutter CM, et al. Estimation of Benefits, Burden, and Harms of Colorectal Cancer Screening Strategies: Modeling Study for the US Preventive Services Task Force. JAMA 2016;315:2595–609.

28.	Ladabaum U, Alvarez-Osorio L, Rösch T, et al. Cost-effectiveness of colorectal cancer screening in Germany: current endoscopic and fecal testing strategies versus plasma methylated Septin 9 DNA. Endosc Int Open 2014;2:E96–E104.

##### **Appendix Table 1** Overview of model parameters

| **A. Proportions of no neoplasm, non-advanced adenoma, advanced adenoma and preclinical CRC at the beginning of simulation^1^** | | | | | |
| --- | --- | --- | --- | --- | --- |
|  |  | **Most advanced finding**  **% (95% confidence interval)** | | | |
| **Sex** |  | **No neoplasm** | **Non-advanced adenoma** | **Advanced adenoma** | **Preclinical colorectal cancer** |
| Men |  | 71.5 (71.3 – 71.7) | 21.7 (21.5 – 21.9) | 6.3 (6.1 – 6.4) | 0.48 (0.45 – 0.52) |
| Women |  | 83.2 (83.0 – 83.3) | 13.2 (13.0 – 13.3) | 3.4 (3.3 – 3.5) | 0.26 (0.24 – 0.29) |
| ^1^ Estimates based on the German screening colonoscopy registry. Extracted and recalculated from reference ^6^ | | | | | |
| **B. Sex- and age-specific annual transition rates between states²** | | | | | |
|  |  | **Annual transition rates**  **% (95% confidence interval)** | | | |
| **Sex** | **Age** | **No neoplasm to non-advanced adenoma** | **Non-advanced adenoma to advanced adenoma** | **Advanced adenoma to preclinical colorectal cancer** | **Preclinical colorectal cancer to clinical colorectal cancer** |
| Men | 50-54 | 3.1 (2.9 – 3.4) | 3.3 (2.8 – 3.9) | 2.6 (2.2 – 3.1) | 17.0 (16.0 – 18.2) |
|  | 55-59 | 3.1 (2.9 – 3.4) | 3.3 (2.8 – 3.9) | 2.6 (2.2 – 3.1) | 17.0 (16.0 – 18.2) |
|  | 60-64 | 3.1 (2.8 – 3.4) | 3.2 (2.6 – 3.7) | 3.1 (2.6 – 3.4) | 18.1 (17.2 – 19.1) |
|  | 65-69 | 3.2 (2.9 – 3.4) | 3.2 (2.6 – 3.7) | 3.8 (3.4 – 4.3) | 20.1 (19.2 – 20.9) |
|  | 70-74 | 2.9 (2.6 – 3.3) | 3.3 (2.6 – 4.0) | 5.1 (4.5 – 5.8) | 19.4 (18.5 – 20.4) |
|  | 75-79 | 2.3 (1.8 – 2.9) | 3.0 (1.9 – 4.2) | 5.2 (4.2 – 6.2) | 19.0 (17.9 – 20.1) |
|  | 80+ | 2.3 (1.8 – 2.9) | 3.0 (1.9 – 4.2) | 5.2 (4.2 – 6.2) | 17.2 (16.0 – 18.8) |
| Women | 50-54 | 1.8 (1.7 – 2.0) | 3.2 (2.6 – 3.8) | 2.5 (2.0 – 2.9) | 20.1 (18.6 – 21.8) |
|  | 55-59 | 1.8 (1.7 – 2.0) | 3.2 (2.6 – 3.8) | 2.5 (2.0 – 2.9) | 20.1 (18.6 – 21.8) |
|  | 60-64 | 2.0 (1.8 – 2.2) | 2.9 (2.2 – 3.4) | 2.7 (2.2 – 3.2) | 21.1 (19.7 – 22.5) |
|  | 65-69 | 2.1 (1.9 – 2.3) | 2.9 (2.3 – 3.5) | 3.8 (3.3 – 4.3) | 20.6 (19.5 – 21.8) |
|  | 70-74 | 2.0 (1.7 – 2.2) | 3.8 (3.0 – 4.6) | 5.0 (4.2 – 5.7) | 19.6 (18.6 – 20.8) |
|  | 75-79 | 1.5 (1.1 – 2.0) | 3.0 (1.7 – 4.4) | 5.6 (4.4 – 6.8) | 18.2 (17.1 – 19.5) |
|  | 80+ | 1.5 (1.1 – 2.0) | 3.0 (1.7 – 4.4) | 5.6 (4.4 – 6.8) | 16.4 (15.3 – 17.8) |
| ² Estimates extracted and recalculated from references ^14–16^ | | | | | |

**Supplementary Table 1** Overview of model parameters (continued)

| **C. Diagnostic performance parameters** | | | | | |
| --- | --- | --- | --- | --- | --- |
|  |  | **Performance (%)** | | | |
| **Test (sex)** | **Parameter** | **No neoplasm** | **Non-advanced adenoma** | **Advanced adenoma** | **Preclinical colorectal cancer** |
| Colonoscopy (both sexes)**³** | Sensitivity | - | 75.0 | 95.0 | 95.0 |
|  | Specificity | 100 | - | - | - |
| FIT (men)^4^ | Sensitivity | - | 15.7 | 31.3 | 80.6 |
|  | Specificity | 91.2 | - | - | - |
| FIT (women)^4^ | Sensitivity | - | 10.7 | 26.3 | 75.6 |
|  | Specificity | 96.2 | - | - | - |
| ³ Estimates based on references ^11,12^  ^4^ Estimates based on references ^20,21^ and ^23–26^. | | | | | |

##### **Appendix Table 2** Annual CRC-specific mortality rates of CRC patients by mode of cancer detection^1^

|  | **Annual CRC-specific mortality rates (%)** | | | |
| --- | --- | --- | --- | --- |
| **Year after diagnosis** | **Screening colonoscopy– detected cases** | | **Symptom-detected cases** | |
|  | **Men** | **Women** | **Men** | **Women** |
| 1 | 4.6 | 3.7 | 19.7 | 20.6 |
| 2 | 2.2 | 1.9 | 9.3 | 10.7 |
| 3 | 2.1 | 1.3 | 8.8 | 7.4 |
| 4 | 1.5 | 0.9 | 6.3 | 4.8 |
| 5 | 1.2 | 0.6 | 5.0 | 3.3 |
| 6 | 0.8 | 0.3 | 3.5 | 1.7 |
| 7 | 0.4 | 0.3 | 1.8 | 1.8 |
| 8 | 0.4 | 0.3 | 1.9 | 1.8 |
| 9 | 0.4 | 0.0 | 1.9 | 0.0 |
| 10 | 0.0 | 0.0 | 0.0 | 0.0 |

^1^ estimates extracted from references ^7,8^

CRC, Colorectal cancer.

##### **Appendix Table 3** Sex- and age-specific general mortality rates

|  | **General mortality rates from age to age +1 (%)**^1^ | |
| --- | --- | --- |
| **Age** | **Men** | **Women** |
| 50 | 0.4 | 0.2 |
| 51 | 0.4 | 0.2 |
| 52 | 0.5 | 0.3 |
| 53 | 0.6 | 0.3 |
| 54 | 0.6 | 0.3 |
| 55 | 0.7 | 0.4 |
| 56 | 0.7 | 0.4 |
| 57 | 0.8 | 0.4 |
| 58 | 0.9 | 0.4 |
| 59 | 1.0 | 0.5 |
| 60 | 1.0 | 0.5 |
| 61 | 1.1 | 0.6 |
| 62 | 1.2 | 0.6 |
| 63 | 1.3 | 0.7 |
| 64 | 1.4 | 0.7 |
| 65 | 1.5 | 0.8 |
| 66 | 1.7 | 0.9 |
| 67 | 1.8 | 0.9 |
| 68 | 1.9 | 1.0 |
| 69 | 2.1 | 1.1 |
| 70 | 2.2 | 1.2 |
| 71 | 2.4 | 1.3 |
| 72 | 2.7 | 1.4 |
| 73 | 3.0 | 1.6 |
| 74 | 3.3 | 1.8 |
| 75 | 3.7 | 2.1 |
| 76 | 4.1 | 2.4 |
| 77 | 4.6 | 2.7 |
| 78 | 5.2 | 3.1 |
| 79 | 5.8 | 3.6 |

*Continued on next page*

**Appendix Table 3.** *Sex- and age-specific general mortality rates (continued)*

|  | **General mortality rates from age to age +1 (%)**^1^ | |
| --- | --- | --- |
| **Age** | **Men** | **Women** |
| 80 | 6.5 | 4.1 |
| 81 | 7.2 | 4.7 |
| 82 | 8.0 | 5.4 |
| 83 | 8.9 | 6.2 |
| 84 | 9.9 | 7.1 |
| 85 | 11.1 | 8.2 |
| 86 | 12.3 | 9.3 |
| 87 | 13.7 | 10.7 |
| 88 | 15.3 | 12.1 |
| 89 | 16.9 | 13.7 |
| 90 | 18.7 | 15.4 |
| 91 | 20.7 | 17.2 |
| 92 | 22.7 | 19.1 |
| 93 | 24.8 | 21.1 |
| 94 | 27.0 | 23.2 |
| 95 | 29.1 | 25.3 |
| 96 | 31.2 | 27.4 |
| 97 | 33.2 | 29.6 |
| 98 | 35.1 | 31.7 |
| 99 | 37.2 | 34.0 |
| 100 | 39.2 | 36.2 |

^1^Estimates were extracted from German population life tables 2010/2012 (reference ^19^).
